# Life Satisfaction in Canada and the Immigrant Experience

**DOI:** 10.1101/2025.06.02.25328814

**Authors:** Sonia S. Anand, Shreni Patel, Scott A. Lear, Trevor J.B. Dummer, Vikki Ho, Jean-Claude Tardif, Jennifer E. Vena, Karleen Schulze, Paul Poirier, Dipika Desai, Matthias G. Friedrich, CAHHM Study Investigators

## Abstract

High-income countries like Canada have the highest self-reported levels of life satisfaction in the world, although little is known regarding the experiences of racialized people and immigrants to these countries. This study investigates the factors that influence subjective well-being among a large national cohort study of 8,063 adults from the Canadian Alliance of Healthy Hearts and Minds cohort study recruited between 2014 and 2018, including a subset of 2,142 immigrants. Measures of demographic, socioeconomic, health, healthcare access, and self-reported ethnicity, from which racialized status was derived, were investigated in relation to self-reported life satisfaction as measured by the validated Cantril ladder score. Among 8,063 adults average age of 58 years, approximately half were women, 18.6% were racialized, and 26.6% were immigrants. Racialized immigrants had significantly lower life satisfaction compared to non-racialized immigrants and Canadian-born persons, whether racialized or not. Multivariable analysis showed that factors associated with higher life satisfaction included older age, male sex, having trusted neighbours, and having a language-concordant family doctor. Factors associated with lower life satisfaction included being racialized, having a higher social disadvantage, poorer cardiovascular health, and being unable to afford prescription medications. Amongst immigrants, those racialized were more likely to report experiencing discrimination based on skin colour and reported lower life satisfaction. Although high income countries like Canada have amongst the highest life satisfaction scores in the world, racialized people, especially immigrants, have lower life satisfaction compared to non-racialized people.

**Key Messages:**

- This study examines associations of demographic, socioeconomic, health, healthcare access that influence self-reported life satisfaction among adults in Canada, particularly among racialized individuals and immigrants.
- Using data from the Canadian Alliance of Healthy Hearts and Minds cohort study, we found racialized immigrants report significantly lower life satisfaction compared to non-racialized immigrants and Canadian-born individuals.
- Despite Canada’s high average life satisfaction, the data suggests there is still a need for equity-focused and culturally tailored healthcare to improve well-being among structurally marginalized groups.

## Introduction

Social determinants such as employment, income, education, housing, and experiences of racism shape health outcomes (1). Although Canada’s 1984 Canada Health Act strives for equal healthcare access, challenges persist due to limited primary care, cultural and language barriers, complex system navigation, and institutional racism (2,3). Research indicates that racialized immigrants are more likely to experience a decline in self-reported health than non-racialized immigrants—a disparity that robust social networks can mitigate (4,5). Furthermore, the traditional "healthy immigrant effect" has weakened, reflecting evolving immigration patterns and an increase in arrivals from conflict zones (6). Life satisfaction, a global measure linked to self-rated health (7–9), is effectively gauged by the validated Cantril Ladder Score, which captures respondents’ current satisfaction and temporal projections (10). Notably, reduced Cantril scores have been associated with chronic diseases, including cardiovascular disease, particularly among immigrant populations (11–14). In this study, we used the Cantril Ladder to assess life satisfaction among participants of the Canadian Alliance for Healthy Hearts and Minds (CAHHM), aiming to elucidate the determinants of life satisfaction among Canadian adults and testing the hypothesis that socially disadvantaged racialized immigrants experience the lowest levels (15).

## Materials and Methods

### Study Design and Participants

This is a cross-sectional analysis of data which were obtained from the CAHHM cohort study, previously described (15). Briefly, the CAHHM recruited adults from the general population between 2014 to 2018 and investigated health behaviours, contextual and socio-cultural factors, chronic disease risk factors and completed a brain and cardiac MRI in all participants. The cohort is currently undergoing repeat examinations 8 years from the baseline assessment. All participants provided written informed consent. Questionnaires were self-administered. The cohort comprises 8,580 individuals (54% women) aged 30 to 78 years from communities across Canada. Ethics approval was obtained from the Hamilton Integrated Research Ethics Board (HiREB #13-255) and all relevant local ethics boards.

### Measurement of Life Satisfaction

The primary outcome variable in this analysis: life satisfaction, was measured using the Cantril ladder score. This psychometrically validated scale offers a quantitative measure of subjective well-being, ranging from 0 (worst possible life) to 10 (best possible life) (10). Cantril Ladder has displayed reliability and validity in diverse adult populations. In a sample of U.S. adults, the ladder had a test-retest correlation of 0.71 over a one month period.(16) The ladder has shown a correlation of 0.68 across five studies of 762 adults with the Satisfaction with Life Scale.(17) Categories of life satisfaction were also examined using predefined cut-offs, including suffering (0 to 4), satisfactory (5 to 6), and thriving (7 to 10).

### Measurement of Sociodemographic Variables/Covariates

Demographic and lifestyle data were collected through standardized questionnaires. The information included demographic variables, cardiovascular risk factors (CVRFs), health behaviours, access to health care, and socioeconomic factors as previously described (15). Ethnicity was self-reported, and from this information, racialized status (referring to non-white individuals) was derived. Physical activity [assessed by short form International Physical Activity Questionnaire (IPAQ-S)], psychological stress, income, employment, and education levels were also self-reported (15). Social disadvantage was derived using an index predictive of CVD (18). Healthcare access was assessed by the Health Services Questionnaire (HSR), which includes questions on access to care, health behaviours, CV risk factors, cardiac diagnostic testing, and information to calculate the non-lab INTERHEART risk score—a validated score for myocardial infarction risk (19,20). Social support was defined as measured using questions about neighbours’ trust and neighbours helping to solve problems (15). Participants in CAHHM who were immigrants completed the Vancouver Index of Acculturation (VIA) (21), which enables scoring to determine an individual’s affinity to their heritage culture and North American culture, years since immigration, and a specific immigrant questionnaire using questions from the Longitudinal Survey of Immigrants to Canada (22).

### Statistical Considerations

All analyses were conducted using SAS (version 9.4), with a statistical significance level set at p < 0.05. Results were reported with corresponding 95% confidence intervals to provide a comprehensive understanding of the precision of estimates. Factors defined above were tested for their association with life satisfaction in univariate analyses, and those with a P <0.20 were entered into a forward stepwise multivariable regression model. Stratified subgroup analyses were conducted to explore potential variations in the impact of determinants on life satisfaction across demographic categories. Interaction terms were incorporated to assess effect modification. For the immigrant subset, additional analyses were conducted to include questions on discrimination.

## Results

### Overall

We stratified participants into four groups based on racialized and immigrant status. The demographic characteristics of all four groups are presented in Table 1. Briefly, we observed the greatest differences in socio-demographic characteristics between non-racialized Canadian-born participants and RI. For example, education level was higher amongst RI compared to non-racialized Canadian-born participants, whereas age, annual income >$100K and CV risk scores were lower among RI compared to non-racialized Canadian-born participants, as were factors such as language-concordance with family doctor, ease of seeing a medical specialist, and neighbourhood support (trust, collective problem solving, and willingness to help each other). Furthermore, there was a higher percentage of RI reported being unable to fill prescriptions due to cost compared to the other groups (Table 1 and Table 2).

**Table 1.** Baseline Demographic Characteristics of Participants Stratified by Immigration and Racialized Status.

|  | Non-racialized<br>Canadian<br>Born | Racialized<br>Canadian<br>Born | Non-racialized<br>Immigrants | Racialized<br>Immigrants<br>(RI) |
| --- | --- | --- | --- | --- |
| Number of participants | 5612 | 309 | 955 | 1187 |
| Female | 3096 (55.2) | 183 (59.2) | 469 (49.1) | 626 (52.7) |
| Age, mean (SD), y | 58.1 (8.6) | 53.9 (9.3) | 60.2 (9.4) | 56.1 (8.9) |
| **Highest Education Attained** |  |  |  |  |
| High school or less | 853 (15.5) | 24 (8.0) | 92 (9.7) | 96 (8.3) |
| College or Trade | 1883 (34.3) | 83 (27.7) | 276 (29.2) | 305 (26.4) |
| University Degree | 2754 (50.2) | 193 (64.3) | 576 (61.0) | 753 (65.3) |
| **Employment** |  |  |  |  |
| Full or part time | 3801 (69.3) | 238 (78.5) | 629 (66.8) | 808 (70.2) |
| Retired | 1307 (23.8) | 46 (15.2) | 246 (26.1) | 244 (21.2) |
| No paid work | 377 (6.9) | 19 (6.3) | 67 (7.1) | 99 (8.6) |
| Living with partner/married | 4080 (74.3) | 213 (70.3) | 737 (78.0) | 911 (79.6) |
| **Average Household Income** |  |  |  |  |
| < \$50,000 | 865 (15.4) | 36 (11.7) | 136 (14.2) | 230 (19.4) |
| \$50,000 - \$74,999 | 944 (16.8) | 48 (15.5) | 180 (18.8) | 196 (16.5) |
| \$75,000 - \$99,999 | 951 (16.9) | 47 (15.2) | 143 (15.0) | 185 (15.6) |
| \$100,000 - \$149,999 | 1289 (23.0) | 61 (19.7) | 214 (22.4) | 223 (18.8) |
| \$150,000 or more | 1144 (20.4) | 85 (27.5) | 210 (22.0) | 217 (18.3) |
| Not captured | 419 (7.5) | 32 (10.4) | 72 (7.5) | 136 (11.5) |

Presented data are n (%) unless otherwise specified.

**Table 2.** Baseline Social Characteristics of Participants Stratified by Immigration and Racialized Status.

|  | **Non-racialized Canadian Born** | **Racialized Canadian Born** | **Non-racialized Immigrants** | **Racialized Immigrants (RI)** |
| --- | --- | --- | --- | --- |
| Number of participants | 5612 | 309 | 955 | 1187 |
| Social disadvantage index, mean (SD) | 1.2 (1.3) | 1.1 (1.3) | 1.3 (1.3) | 1.3 (1.3) |
| **Social disadvantage** |  |  |  |  |
| Low disadvantage | 3050 (54.3) | 182 (58.9) | 507 (53.1) | 625 (52.7) |
| Mod disadvantage | 1801 (32.1) | 78 (25.2) | 321 (33.6) | 348 (29.3) |
| High disadvantage | 331 (5.9) | 17 (5.5) | 52 (5.4) | 71 (6.0) |
| Missing value | 430 (7.7) | 32 (10.4) | 75 (7.9) | 143 (12.0) |
| If a problem, neighbours work together to deal with it (agree) | 4224 (75.6) | 228 (74.0) | 736 (77.8) | 848 (71.7) |
| Neighbours trust each other | 5224 (93.5) | 276 (89.6) | 880 (93.0) | 1044 (88.3) |
| Neighbours willing to help their neighbours | 5040 (90.1) | 271 (87.7) | 856 (90.1) | 1029 (86.9) |
| INTERHEART risk score, mean (SD) | 10.5 (6.0) | 9.5 (5.6) | 10.2 (5.9) | 9.6 (5.4) |
| **INTERHEART risk score** |  |  |  |  |
| Low | 2689 (48.1) | 157 (50.8) | 484 (50.8) | 637 (53.8) |
| Moderate | 1793 (32.1) | 106 (34.3) | 307 (32.2) | 387 (32.7) |
| High | 1108 (19.8) | 46 (14.9) | 161 (16.9) | 159 (13.4) |
| **Access to Health Services** |  |  |  |  |
| Have a regular primary care provider | 5317 (94.8) | 293 (94.8) | 910 (95.4) | 1132 (95.5) |
| Visited primary care provider in past 12 months |  |  |  |  |
| Yes | 4803 (85.6) | 243 (78.6) | 824 (86.4) | 1057 (89.2) |
| No or do not know | 512 (9.1) | 50 (16.2) | 86 (9.0) | 75 (6.3) |
| Do not have a primary care provider | 293 (5.2) | 16 (5.2) | 44 (4.6) | 53 (4.5) |
| Speak same language as primary care provider |  |  |  |  |
| Yes | 5283 (94.3) | 292 (94.5) | 893 (93.6) | 1036 (87.5) |
| No | 28 (0.5) | 1 (0.3) | 17 (1.8) | 95 (8.0) |
| Do not have a primary care provider | 293 (5.2) | 16 (5.2) | 44 (4.6) | 53 (4.5) |
| Went to Emergency in past 12 months | 907 (16.2) | 38 (12.3) | 139 (14.6) | 150 (12.7) |
| Experience any difficulties getting the routine or ongoing care | 618 (11.0) | 31 (10.0) | 104 (10.9) | 123 (10.4) |
| Difficulties getting specialist care in past 12 months |  |  |  |  |
| Yes | 501 (8.9) | 20 (6.5) | 97 (10.2) | 134 (11.3) |
| No or do not know | 2582 (46.1) | 127 (41.1) | 460 (48.4) | 497 (41.9) |
| Did not need specialist | 2523 (45.0) | 162 (52.4) | 394 (41.4) | 554 (46.8) |
| Did not fill prescription because of the cost | 144 (2.6) | 13 (4.2) | 27 (2.8) | 64 (5.4) |
| Cantril Ladder Score | 7.2 (1.4) | 7.2 (1.5) | 7.2 (1.5) | 6.6 (1.6) |
| **Cantril Categories** |  |  |  |  |
| Suffering (1-4) | 243 (4.3) | 17 (5.5) | 42 (4.4) | 105 (8.8) |
| Struggling (5-6) | 1270 (22.6) | 61 (19.7) | 229 (24.0) | 402 (33.9) |
| Thriving (7-10) | 4099 (73.0) | 231 (74.8) | 684 (71.6) | 680 (57.3) |

Presented data are n (%) unless otherwise specified.

The life satisfaction scores were lowest among RI compared to all other groups, using non-racialized Canadian-born participants as the reference group (6.6 ± 1.6 vs. 7.2 ± 1.4), P<0.0001). These differences between RI and non-racialized Canadian-born participants persisted after adjustment for years of living in Canada [6.8 (95% CI: 6.7-6.9) vs. 7.2 (95% CI: 7.1-7.2)] (Figure 1). RI compared to non-racialized Canadian-born participants were less likely to report a ladder score in the thriving range of 7 to 10 (57.3 vs. 73.0%), and more likely to report scores in the struggling range of 5 to 6 (33.9 vs. 22.6%) or in the suffering range of 0 to 4 (8.8 vs. 4.3%), respectively (Table 2).

**Figure 1.**
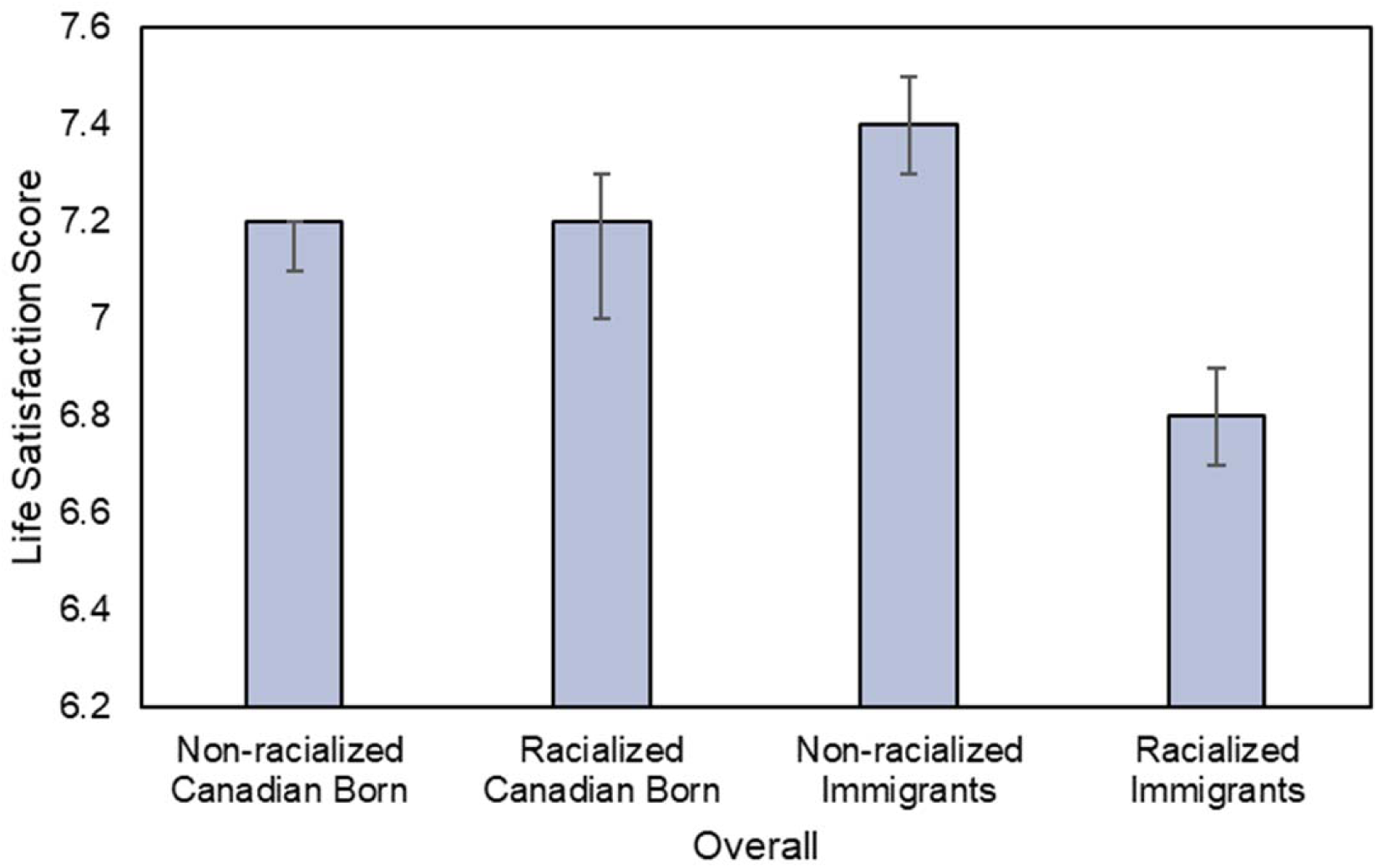
Life satisfaction scores across demographic groups (adjusted for years in Canada). P for differences between groups <0.0001.

The factors associated with life satisfaction in all participants are shown in Figure 2a. Briefly, in order of magnitude, factors associated with lower life satisfaction included not being able to fill a prescription due to cost, social disadvantage, being an RI, female sex, the presence of CVRFs, and having had an emergency room visit in the last 12 months. Factors associated with improved life satisfaction included having trusted neighbours, having a language-concordant primary care doctor, having neighbours to help solve problems, and increasing age (Figure 2a and Table 3a).

**Figure 2.**
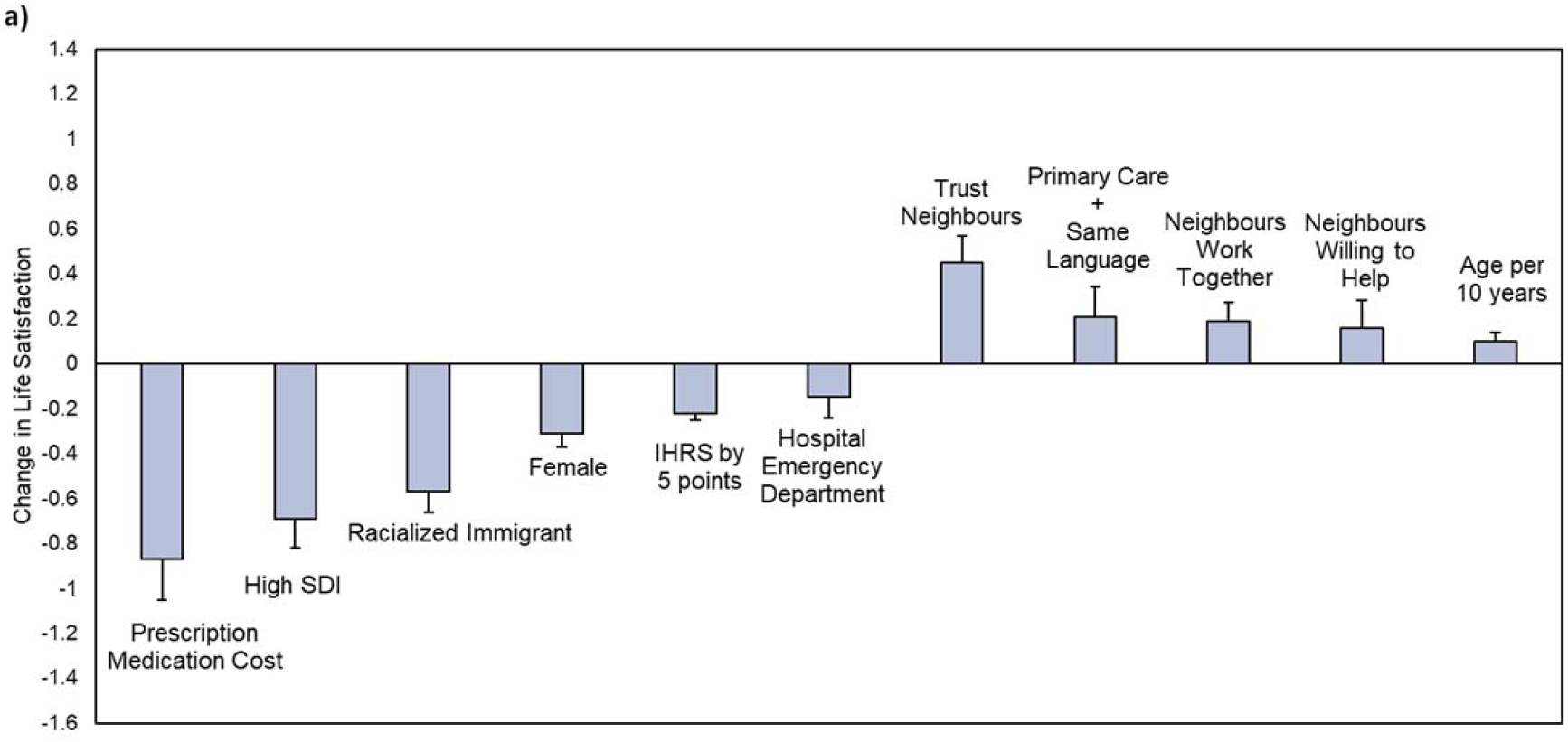
(a) Determinants of life satisfaction scores in the overall cohort. *SDI = Social Disadvantage; IHRS = INTERHEART Risk Score; Discrim = Discrimination

**Figure 2.**
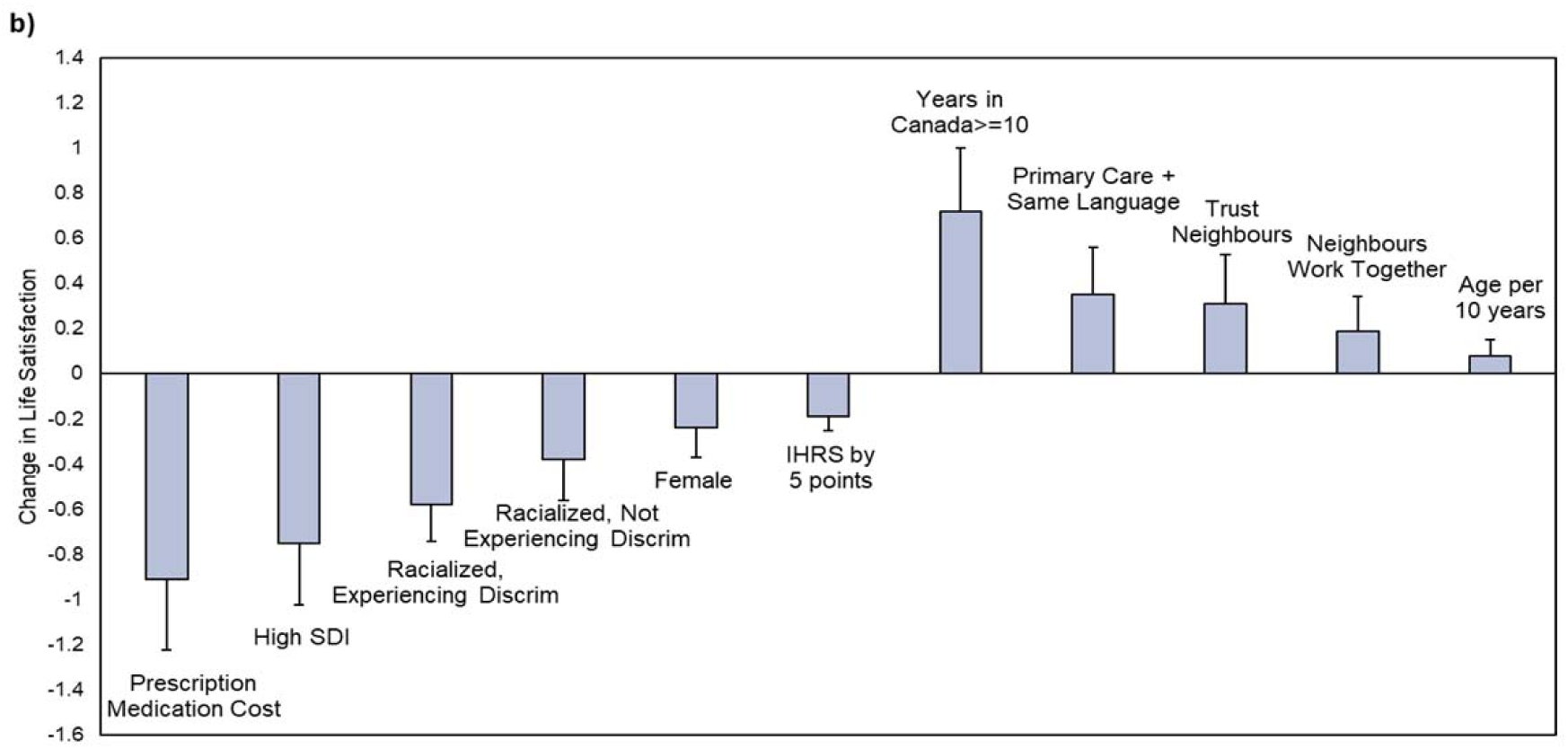
(b) Determinants of life satisfaction scores in the immigrant cohort. *SDI = Social Disadvantage; IHRS = INTERHEART Risk Score; Discrim = Discrimination

**Table 3a.**
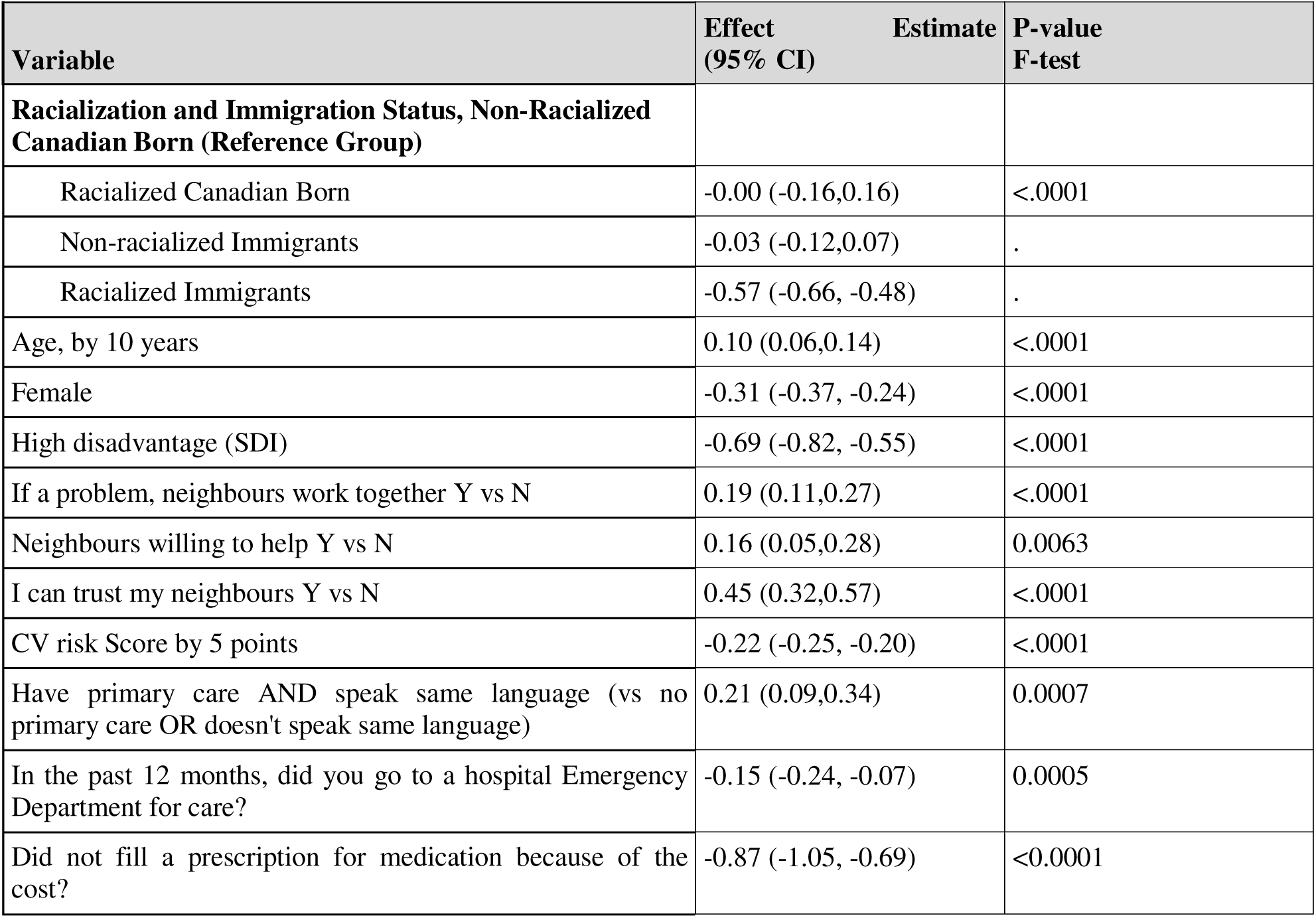
Predictors of the Cantril Score (continuous) in all participants using a Multivariable Model (N=7970)

**Table 3b.** Predictors of Cantril Score (continuous) in Immigrant Subpopulation using a Multivariable Model (N=2013)

| Variable | Effect<br>95% CI | P-value<br>F-test |
| --- | --- | --- |
| **Racialization and Discrimination, Non-Racialized Not Experiencing Discrimination (Reference Group)** |  |  |
| Racialized Not Experiencing Discrimination | -0.38 (-0.56, -0.20) | <.0001 |
| Non-racialized Experiencing Discrimination | -0.01 (-0.22,0.21) | . |
| Racialized Experiencing Discrimination | -0.58 (-0.74, -0.43) | . |
| Age, by 10 years | 0.08 (0.00,0.15) | 0.0389 |
| Female | -0.24 (-0.37, -0.11) | 0.0003 |
| High disadvantage (SDI) | -0.75 (-1.02, -0.48) | <.0001 |
| If a problem, neighbours work together Y vs N | 0.19 (0.04,0.34) | 0.0117 |
| I can trust my neighbours Y vs N | 0.31 (0.10,0.53) | 0.0049 |
| CV risk Score by 5 points | -0.19 (-0.25, -0.13) | <.0001 |
| Have primary care AND speak same lang (vs no primary care OR doesn't speak same language) | 0.35 (0.14,0.56) | 0.0012 |
| Did not fill a prescription for medication because of the cost? | -0.91 (-1.22, -0.61) | <.0001 |
| Years in Canada >=10 | 0.72 (0.43,1.00) | <.0001 |

### Immigrant subset

Amongst immigrants, questions regarding discrimination were collected as part of the acculturation questionnaire (Table 4). Among 2,142 who completed these questions, RI reported experiencing more discrimination compared to non-racialized immigrants (64 vs. 27%, respectively). The discrimination based on skin colour (as a proportion of the ethnic group) was largely driven by the experience of Black participants (i.e., 67% in Black people vs. 37% South Asians, 34% Chinese, and 30% in the other immigrant category). The most reported reasons for discrimination for both racialized and non-racialized are shown in Figure 3.

**Figure 3.**
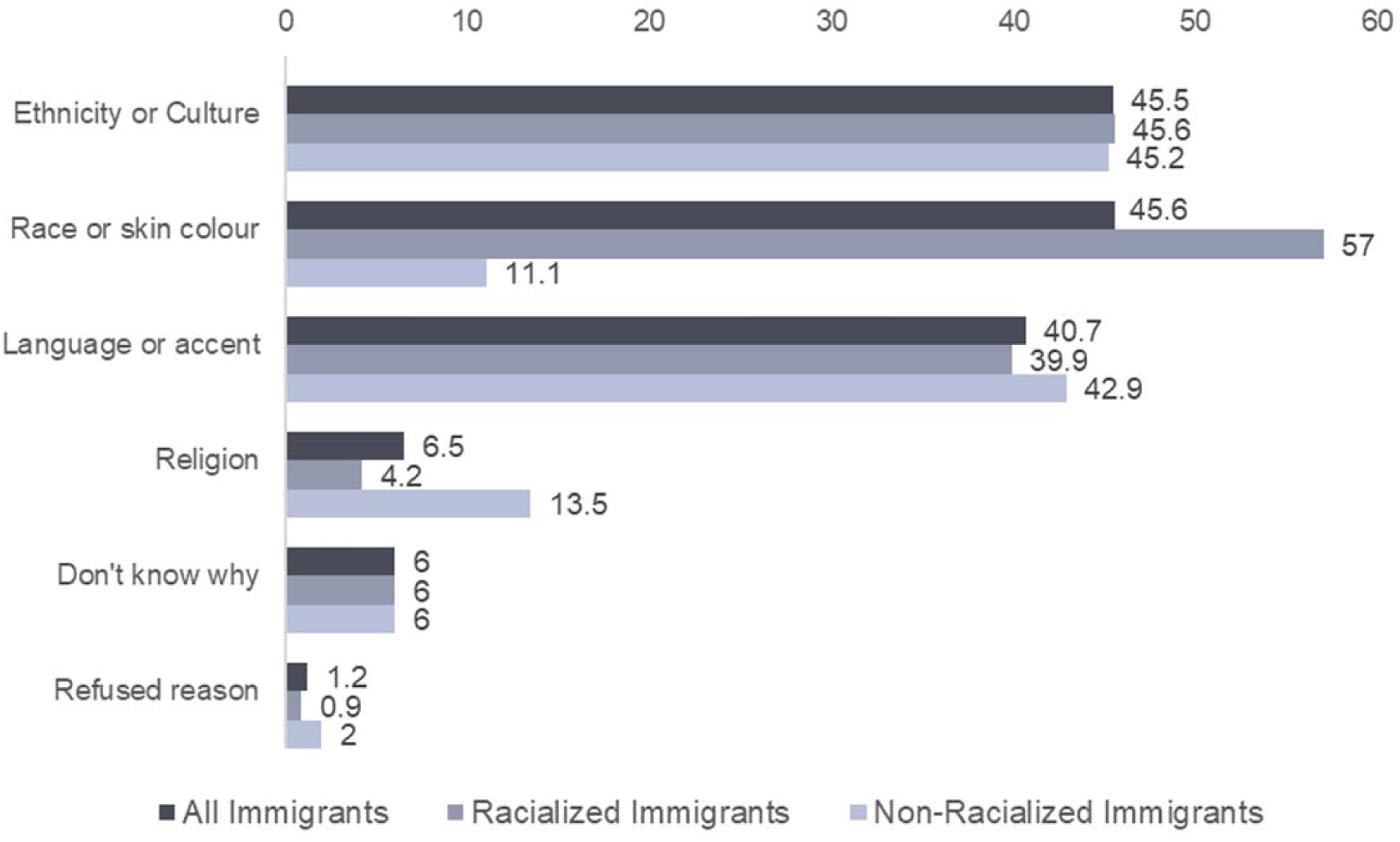
Commonly reported reasons for discrimination among Immigrants.

**Table 4.** Baseline Characteristics for Immigrant Subpopulation Specific Measures.

|  | All Immigrants to Canada | Racialized Immigrants | Non-Racialized Immigrants |
| --- | --- | --- | --- |
| Number of participants | 2142 | 1187 | 955 |
| Have you experienced discrimination or been treated unfairly by others | 1013 (47.5) | 761 (64.3)* | 252 (26.5) |
| Reasons<sup>1</sup> for experienced discrimination |  |  |  |
| Ethnicity or Culture | 461 (45.5) | 347 (45.6) | 114 (45.2) |
| Race or skin colour | 462 (45.6) | 434 (57.0)* | 28 (11.1) |
| Language or accent | 412 (40.7) | 304 (39.9) | 108 (42.9) |
| Religion | 66 (6.5) | 32 (4.2)* | 34 (13.5) |
| Refused reason | 12 (1.2) | 7 (0.9) | 5 (2.0) |
| Don't know why | 61 (6.0) | 46 (6.0) | 15 (6.0) |
| VIA, mean (SD) |  |  |  |
| VIA-Heritage affinity | 6.4 (1.3) | 6.4 (1.3)* | 6.3 (1.4) |
| VIA-North American affinity | 6.5 (1.3) | 6.2 (1.2)* | 6.9 (1.2) |
| **How long have you lived in Canada, mean (SD)** | 34.1 (16.0) | 29.5 (13.5)* | 39.9 (16.9) |
| < 10 years | 112 (5.2) | 86 (7.2) | 26 (2.7) |
| 10 - 19 years | 374 (17.5) | 232 (19.5) | 142 (14.9) |
| 20 - 29 years | 436 (20.4) | 321 (27.0) | 115 (12.0) |
| 30 - 39 years | 316 (14.8) | 197 (16.6) | 119 (12.5) |
| 40 - 49 years | 556 (26.0) | 289 (24.3) | 267 (28.0) |
| 50 - 59 years | 205 (9.6) | 51 (4.3) | 154 (16.1) |
| 60+ years | 143 (6.7) | 11 (0.9) | 132 (13.8) |

Presented data are n (%) unless otherwise specified.
<sup>1</sup>Multiple responses possible.
\*p<.05 comparing racialized and non-racialized immigrants.
VIA = Vancouver Index of Acculturation

Other differences between RI vs. non-RI included their lower affinity to North American culture and their shorter duration of time living in Canada, which was, on average 10 years less. (Figure 4)

**Figure 4.**
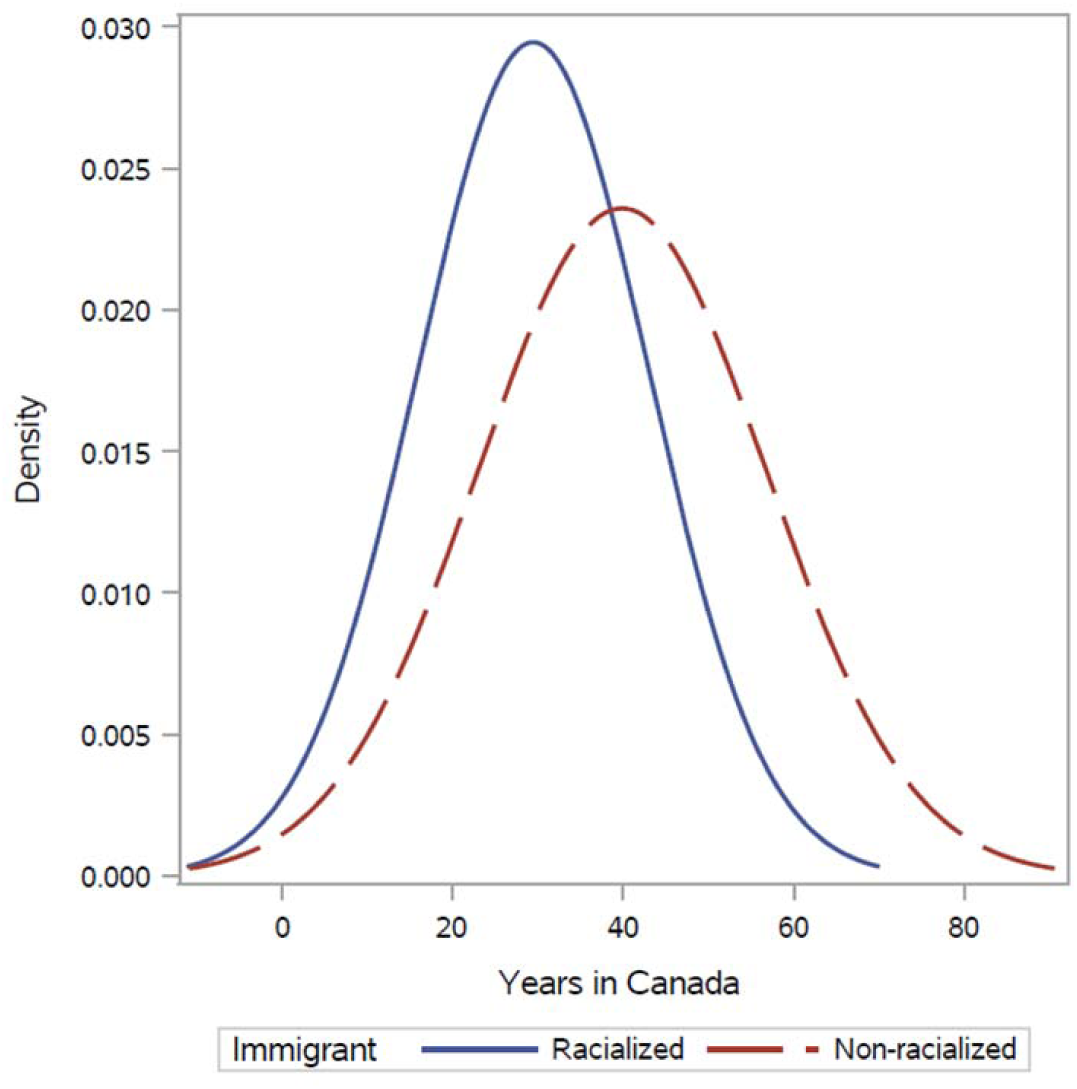
Immigrant years in Canada by racialized status.

Amongst immigrants in the multivariate regression model, factors associated with lower life satisfaction in order of magnitude included: not being able to fill a prescription due to cost, high social disadvantage, experiencing discrimination, being racialized (even if they did not report discrimination), female sex, and having CVRFs. Factors associated with higher life satisfaction included years lived in Canada, having a language-concordant primary care doctor, having trusted neighbours and older age (Figure 2b and Table 3b).

## Discussion

Our cross-sectional study of 8,063 adults finds racialized immigrants (RI) have significantly lower life satisfaction scores than other groups, particularly those experiencing discrimination. Low life satisfaction correlates with factors including inability to afford prescriptions, higher social disadvantage, being female, and more cardiovascular (CV) risk factors. Protective factors include social support, access to a language-concordant family doctor, and longer duration of residence in Canada.

The strongest determinant of lower life satisfaction was the inability to afford prescriptions, reflecting a broader issue of financial hardship and uneven access to healthcare. Canada’s fragmented medication coverage further exacerbates this issue. Structural inequalities persist, particularly for racialized groups, where both financial constraints and healthcare access contribute to lower satisfaction. Addressing these disparities through national pharmacare and improving language-concordant care may help mitigate these issues. Social disadvantage, including employment and housing instability, remains a powerful determinant in life satisfaction, emphasizing the need for targeted policy interventions. Strengths of our study include a large, diverse sample and the use of the validated Cantril Ladder to assess life satisfaction. Limitations include the cross-sectional nature of the analysis, preventing causal conclusions. Future prospective analyses could further elucidate the relationship between life satisfaction and long-term health outcomes.

This cross-sectional analysis of 8,063 Canadian adults reveals that while overall life satisfaction is high, racialized immigrants (RI) report significantly lower scores—especially when facing discrimination. Key factors linked to lower life satisfaction include financial hardship (notably, the inability to pay for prescription medications), higher social disadvantage, female gender, and increased cardiovascular risk factors. In contrast, protective factors such as trustworthy neighbors, a language-concordant primary care physician, older age, and longer residence in Canada for immigrants buffer these negative effects.

Health status and access play a critical role; the inability to afford prescription medications— indicative of financial strain leading to medication rationing—was the strongest determinant of lower life satisfaction (23). This issue is compounded by provincial disparities in prescription coverage and Canada’s lack of a national pharmacare plan, despite being the only G7 country without one (24,25). Additional adverse factors include recent emergency room visits and cardiovascular risk burdens, while language-concordant care is associated with improved outcomes.

Social disadvantage—a composite of employment, income, and marital status—underscores the influence of structural racism on health. Our findings, consistent with prior studies (26–28), indicate that racialized individuals experience higher levels of discrimination in healthcare, contributing to their lower life satisfaction. There is some evidence that perceived racial discrimination is associated with measurable brain changes on MRI—particularly in regions involved in stress, vigilance, and threat detection (29). This will be the focus of future analyses. Moreover, our analysis shows that immigrants tend to have a stronger affinity for their heritage culture, whereas Canadian-born individuals exhibit a greater affinity for Canadian culture. This cultural alignment may further influence their overall well-being.

Strengths of this study include its large, multi-ethnic sample and the use of the validated Cantril Ladder to assess life satisfaction. However, the cross-sectional design limits causal interpretations, and the CAHHM cohort’s demographic profile—older, more highly educated, and with fewer cardiovascular risk factors—might underestimate the broader impact of these determinants. Future longitudinal analyses will be essential to better understand the temporal relationships between factors such as perceived discrimination, brain MRI changes, and life satisfaction.

## Conclusions

Our comprehensive analysis of the determinants of life satisfaction in Canada shows the complex interactions between various social, demographic and healthcare factors, which offers valuable insights into the immigrant experience in Canada. Our findings contribute to the evidence base necessary for informed policy formulation aimed at enhancing the overall life satisfaction of diverse communities in Canada.

## Ethics approval

Ethics approval was obtained from the Hamilton Integrated Research Ethics Board (HiREB #13-255) and all relevant local ethics boards. All participants provided written informed consent.

## Data availability

The data underlying this article will be shared on reasonable request to the corresponding author.

## Author contributions

SSA developed the idea and drafted the manuscript, KS led the statistical analysis, DD coordinated the study, SP assisted with writing, editing, and creation of the figure. All others edited the manuscript.

## Funding

CAHHM was funded by the Canadian Partnership Against Cancer (CPAC), Heart and Stroke Foundation of Canada (HSF-Canada), and the Canadian Institutes of Health Research (CIHR). Financial contributions were also received from the Population Health Research Institute and CIHR Foundation Grant no. FDN-143255 to S.S.A.; FDN-143313 to J.V.T.; and FDN 154317 to E.E.S. In-kind contributions from A.R.M. and S.E.B. from Sunnybrook Hospital, Toronto for MRI reading costs, and Bayer AG for provision of IV contrast. The Canadian Partnership for Tomorrow’s Health is supported by the Canadian Partnership Against Cancer, BC Cancer, Genome Quebec, Centre de recherche CHU Sainte-Justine, Ontario Institute for Cancer Research, Alberta Health, Alberta Cancer Foundation, Alberta Health Services, and Dalhousie University. The PURE Study was funded by multiple sources. The Montreal Heart Institute Biobank is funded by Mr André Desmarais and Mrs France Chrétien-Desmarais and the Montreal Heart Institute Foundation. S.S.A. was supported by the Heart and Stroke Foundation Chair in Population Health. S.A.L is supported by the Pfizer/Heart & Stroke Foundation Chair in Cardiovascular Prevention Research at St. Paul’s Hospital. P.A. was supported by a Ministry of Research and Innovation of Ontario Investigator Award. S.E.B. was supported by the Hurvitz Brain Sciences Research Program, Sunnybrook Research Institute, and the Department of Medicine, Sunnybrook Health Sciences Centre, University of Toronto. E.L. was supported by the Laval University Chair of Research & Innovation in Cardiovascular Imaging and the Fonds de recherche du Québec—Santé. J.-C.T. holds the Tier 1 Canada Research Chair in translational and personalized medicine and the Université de Montréal Pfizer endowed research chair in atherosclerosis. CG is supported by grants from the Swiss National Science Foundation (SNSF, # PP00P3_163892 and # PP00P3_190074), the Olga Mayenfisch Foundation, Switzerland, the OPO Foundation, Switzerland, the Novartis Foundation, Switzerland, the Swissheart Foundation, the Helmut Horten Foundation, Switzerland, the University Hospital Zurich (USZ) Foundation, the Iten-Kohaut Foundation, Switzerland, and the EMDO Foundation, Switzerland.

Some of the data used in this research were made available by the Canadian Partnership for Tomorrow’s Health along with BC Generations Project, Alberta’s Tomorrow Project, Ontario Health Study, CARTaGENE, and the Atlantic PATH. Data were harmonized by Maelstrom Research and access policies and procedures were developed by the Centre of Genomics and Policy in collaboration with the Cohorts.

## Acknowledgements

**Steering Committee of CAHHM:** S. Anand (Chair)*, M.G. Friedrich (Co-Chair), J. Tu (Co-Chair), P Awadalla (OHS), T. Dummer (BCGP), J. Vena (ATP), G. Lettre, V. Ho (CaG), J. Hicks (APATH), J-C. Tardif (MHI Biobank), K. Teo, S. Yusuf (PURE-Central), B-M. Knoppers (ELSI).

**Project Office Staff at Population Health Research Institute (PHRI):** D. Desai

**Statistics/Biometrics Programmers Team at PHRI:** K. Schulze, S. Bangdiwala, C. Ramasundarahettige, K. Ramakrishnana

**Central Operations Leads:** D. Desai (PHRI), Sherry Zafar, Andrea Rogge

**Cohort Operations Research Personnel:** K. McDonald (OHS), N. Noisel (CaG), J. Chu (BCGP), J. Hicks (APATH), H. Whelan (ATP), S. Rangarajan (PURE), D. Busseuil (MHI Biobank)

**Site Investigators and Staff:** (112) J. Leipsic, S. Lear, V. de Jong; (306) M. Noseworthy, K. Teo, E. Ramezani, N. Konyer; (402) P. Poirier, A-S. Bourlaud, E. Larose, K. Bibeau; (512) J. Leipsic, S. Lear, V. de Jong; (609) E. Smith, R. Frayne, A. Charlton, R Sekhon; (703) A. Moody, V. Thayalasuthan; (704) A. Kripalani, G Leung; (706) M. Noseworthy, S. Anand, R. de Souza, N. Konyer, S. Zafar; (707) G. Paraga, L. Reid; (714) A. Dick, F. Ahmad; (799) D. Kelton, H. Shah; (801) F. Marcotte, H. Poiffaut; (802) M. Friedrich, J. Lebel; (817) E. Larose, K. Bibeau; (913) L. Parker, D. Thompson, J. Hicks; (1001) J-C. Tardif, H. Poiffaut; (1103) J. Tu, K. Chan, A. Moody, V. Thayalasuthan;

## MRI Working Group and Core Lab Investigators/Staff

**Chair:** M.G. Friedrich; Brain Core Lab: E. Smith, C. McCreary, S. E. Black, C. Scott, S. Batool, F. Gao; Carotid Core Lab: A. Moody, V. Thayalasuthan; Abdomen: E. Larose, K. Bibeau,

**Cardiac:** F. Marcotte, F. Henriques, T. Teixeira.

**Contextual Working Group:** R. de Souza, S. Anand, G. Booth, J. Brook, D. Corsi, L. Gauvin, S. Lear, F. Razak, S.V. Subramanian, J. Tu.

**CAHHM Founding Advisory Group:** Jean Rouleau, Pierre Boyle, Caroline Wong, Eldon Smith

## Conflicts of Interest

The authors have no conflicts of interest to disclose.

## Figures

**Supplemental Figure 1.**
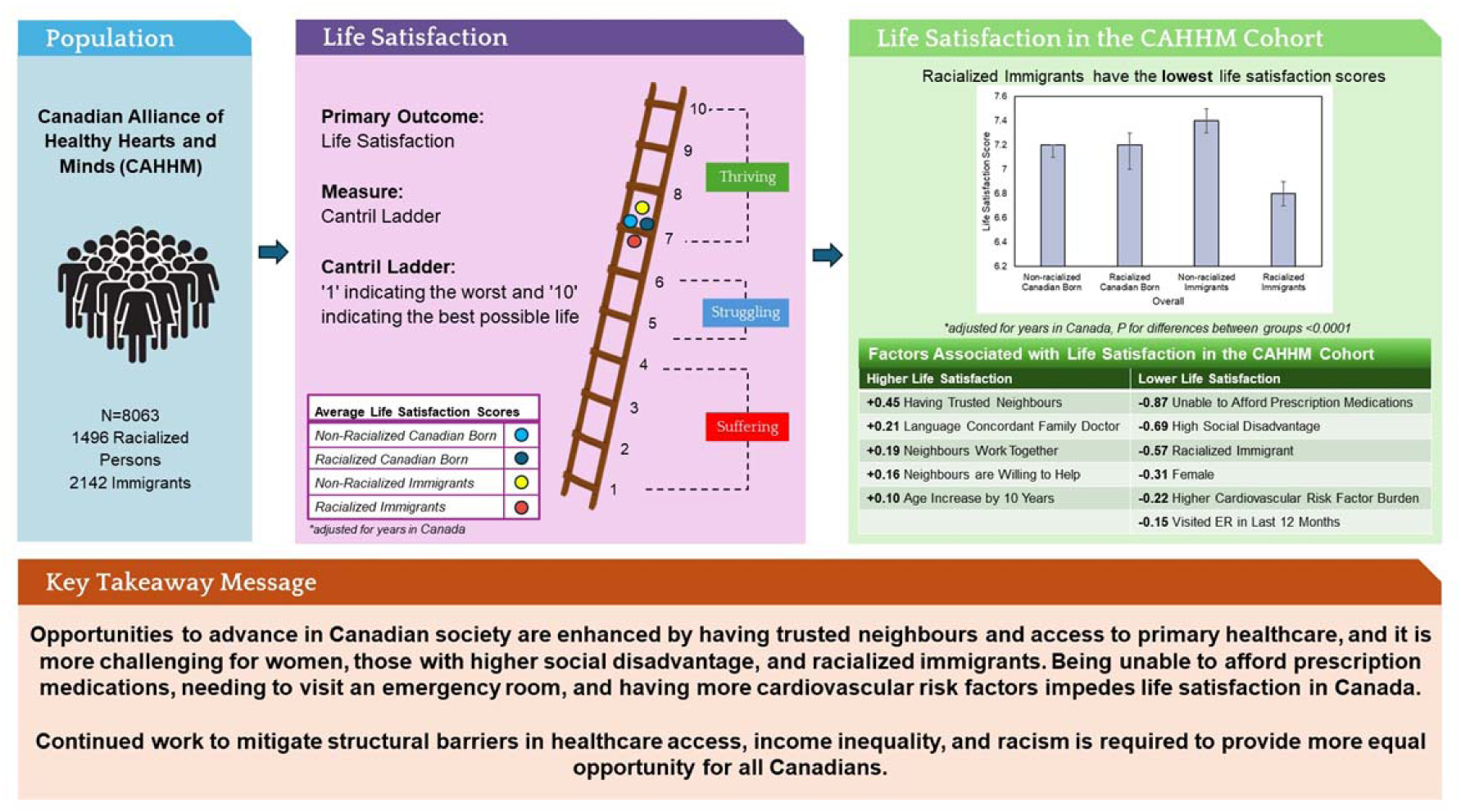
Graphical Abstract.

## Notes

### Competing Interest Statement

The authors have declared no competing interest.

### Author Declarations

Ethics Committee of Hamilton Integrated Research Ethics Board gave ethical approval for this work (HiREB #13-255).

